# Spatio-Temporal Dynamics and Climate Change Scenarios Forecast of Dengue Incidence in Brazil

**DOI:** 10.1101/2024.07.22.24310334

**Authors:** Patricia Marques Moralejo Bermudi, Francisco Chiaravalloti Neto, Marta Blangiardo, Raquel Gardini Sanches Palasio, Anderson de Oliveira, Monica Pirani

## Abstract

Dengue is a neglected tropical vector-borne disease that is spreading rapidly and increasing worldwide. Climate change has been considered one of the primary factors contributing to this escalation. In Brazil, a vast and heterogeneous country, each epidemic has set new records, with the disease currently spreading widely across the country. In this study, we used predictive modeling techniques, grounded on a Bayesian hierarchical framework, to forecast the spatio-temporal dynamics of dengue in Brazil under future climate scenarios. We used monthly historical data on dengue incidence from 2000 to 2021, collected over 557 Brazilian microregions, to learn about its association with population, environmental, climatic, and socioeconomic conditions. By integrating projections of precipitation, minimum temperature and population based on different Shared Socioeconomic Pathway scenarios, we obtained spatially resolved projections of dengue incidence for 2021-2040 and 2041-2060. The model’s predictive performance was ensured via robust cross-validation. We observed an increase in dengue incidence with rising precipitation, urban infrastructure, and Normalized Difference Water Index, and a decrease with increas-ing elevation and deprivation index. Minimum temperature exhibited a nonlinear and positive association behavior. Our forecasts show great regional variability. In the south region, some traditionally non-endemic microregions are expected to see a clear increase in dengue incidence rate. Conversely, other regions show geographic variability of patterns, suggesting that in some cases elevated temperatures may exceed the viable threshold for dengue transmission. Investing in reducing greenhouse gas emissions and implementing measures to address the rising incidence of dengue due to climate change is crucial.

## 1 Introduction

Dengue is considered the most important mosquitoborne viral disease in humans, caused by arboviruses with four serotypes [1]. Its symptoms can often overlap with other diseases, including COVID-19 [2]. This makes its diagnosis challenging, especially in re-gions where it is endemic, which include several tropical and subtropical countries worldwide. Globally, there has been a significant increase in dengue incidence over time, with an extension in the geographical range of transmission [3–5]. In the Americas, the pattern observed is cyclic, with outbreaks every three to five years [6].

In Brazil, the first reports of dengue date back to the end of the 19th century [1]. Despite significant efforts in surveillance and control measures, including a government investment of over half a billion dollars annually in mosquito control, there has been no reduction in vector density that could effectively limit or decrease the sustained spread of dengue. The disease remains a pressing public health concern [7]. Recently, in 2024, dengue cases and deaths reached a historic record with over 6.3 million probable cases and more than 4,700 confirmed deaths [8].

The increase in the incidence of dengue fever, both globally and in Brazil, is increasingly being attributed to global warming and climate change. Recent findings from the Intergovernmental Panel on Climate Change (IPCC) highlight an alarming trend where human activities are driving an irreversible rise in global temperatures [9].

Climatic conditions significantly influence various aspects of dengue transmission, including the virus (e.g., incubation period), the vector (e.g., accelerated development rates), and human behavior (e.g., water storage during drought periods) [10, 11].

Other contributing factors include sanitation, human mobility, deficiencies in mosquito control measures, population growth, urbanization, and improved disease reporting capabilities [12].

In Brazil, the current strategic tools employed to identify areas at risk and address dengue epidemics primarily focus on entomological surveillance, relying on indicators that assess the immature stages of *Aedes aegypti* (Linnaeus, 1762) [13] which is the primary vector. However, numerous authors have criticized the use of these indicators, which are recommended by the Ministry of Health, as they do not provide estimates of adult mosquito abundance or direct assessments of dengue transmission risk. In practice, a gap exists between traditional entomological surveillance measures and the implementation of vector control interventions, leading to indiscriminate control efforts that fail to prioritize high-risk areas [14–16]

Furthermore, the direct correlation between control measures and the reduction in dengue cases remains inadequately supported, necessitating further research to explore this relationship. It is noteworthy that in Brazil, there have been no significant changes in vector control activities since the beginning of the first dengue epidemics, after the reintroduction of the vector in Brazil in the 1970s [17]. It is also worth mentioning that, despite promising advances in vaccine research, a vaccine has not yet been implemented for the entire population [18].

Presenting a significant public health challenge, dengue is currently categorized as a neglected tropical disease (NTD), and its control efforts are in alignment with the Sustainable Development Goals (SDGs) established by the United Nations. Specifically, SDG Target 3.3 aims to eliminate NTDs by 2030 [19]. In Brazil, cost estimates have ranged from 516.79 million USD (2009) to 1,988.3 million USD (2013) [20]. The burden extends beyond economic considerations, encompassing societal impacts as well, and it appears to be on the rise, potentially underestimated [21].

This paper presents a modelling study of dengue incidence and its main determinants in Brazil, accounting for spatial and temporal dependencies, and use the approach to project dengue incidence to 2021-2060 based on different scenarios of greenhouse gas emissions.

## Results

The findings in this section are derived from a Bayesian hierarchical mixed effect model with spatiotemporal random fields, assuming a Poisson distribution for the monthly counts of dengue cases reported across 557 Brazilian microregions (which are legally defined areas consisting of groups of municipalities) from 2000 to 2021. We selected seven predictors to explain the spatio-temporal variability of the dengue incidence rates (per 100,000 people), including minimum temperature, precipitation, percentage of ur-ban infrastructure, the Normalized Difference Water Index (NDWI), elevation, seasonal period and the Brazilian deprivation index (BDI).

The effects of the covariates on the dengue incidence rate (per 100,000 people) are shown in Table 1. These results, summarised by the posterior mean and the 95% credible intervals (CI), are presented on the incidence rate ratio (IRR) scale, for a one standard deviation increase in each covariate value (since the covariates were standardized before being entered into the spatio-temporal model). They reveal a positive association with lagged effect (two months) of precipitation, with one standard deviation increase corresponding to a 4% (95% CI: 4-5%) escalation in the incidence rate of dengue.

**Table 1:** Covariate effects, described by the posterior means and 95% credible intervals (CI), on the dengue incidence rate (per 100,000 people) in Brazil from 2000 to 2021.

| Covariates | IRR | 95% CI |
| --- | --- | --- |
| Precipitation (2-month lag) | 1.04 | (1.03-1.05) |
| % Urban Infrastructure | 1.08 | (1.03-1.12) |
| NDWI | 1.08 | (1.06-1.09) |
| Elevation | 0.69 | (0.65-0.73) |
| Seasons: |  |  |
| Winter (ref.) | 1 | - |
| Spring | 0.90 | (0.74-1.12) |
| Summer | 3.20 | (2.58-3.93) |
| Autumn | 3.07 | (2.48-3.75) |
| Deprivation Index (BDI): |  |  |
| BDI < -0.7 (ref.) | 1 | - |
| -0.7 ≤ BDI < 0.7 | 0.71 | (0.64-0.78) |
| BDI ≥ 0.7 | 0.46 | (0.40-0.53) |

Similarly, one standard deviation increase in NDWI, % urban infrastructure, and elevation corresponds to a 8% (95% CI: 6-9%), 8% (95% CI: 3-12%) increase, and a 31% (95% CI: 27-35%) decrease, respectively, in the dengue incidence rate.

We also found significant relationships between dengue incidence and the seasons of the year and the deprivation index. In detail, we estimated a higher incidence rate in summer and autumn compared to winter and a negative association of dengue incidence rate with the deprivation index.

Figure 1 displays a nonlinear relationship between dengue incidence rate and minimum temperature. It is noted that the temperature begins to show a positive relationship with the incidence of dengue around 15 °C, and as the temperature increases, a downward trend in this association is observed, although it remains largely positive and significant.

**Figure 1:**
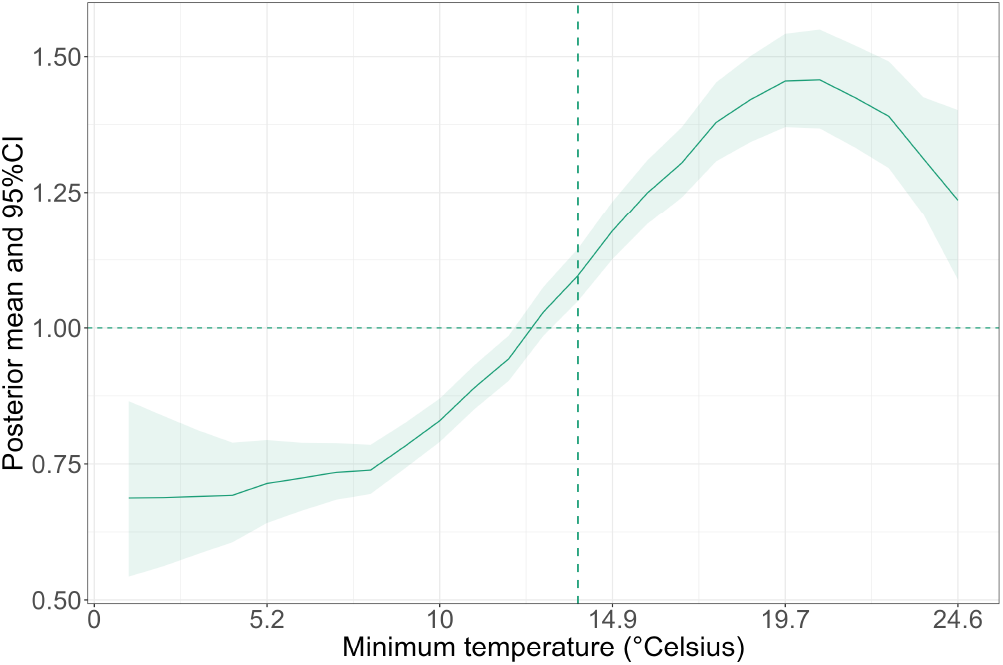
Effect of minimum temperature, described by the posterior mean (green solid line) and 95% credible intervals (CI; green ribbon) on the dengue incidence rate (per 100,000 people) in Brazil from 2000 to 2021

Figure 2 presents the results of cross-validation for the selected model. In each iteration of the training and testing rounds, data from one month was excluded from all microregions within a specific region of Brazil in the training set, while the remaining data was used for prediction. This process was repeated for all twelve months across all five regions of Brazil. We observed high correlation between the observed and predicted values, as well as MAE values lower than the average of the cases. These results indicate that the model is effective in making predictions considering different months and regions of Brazil.

**Figure 2:**
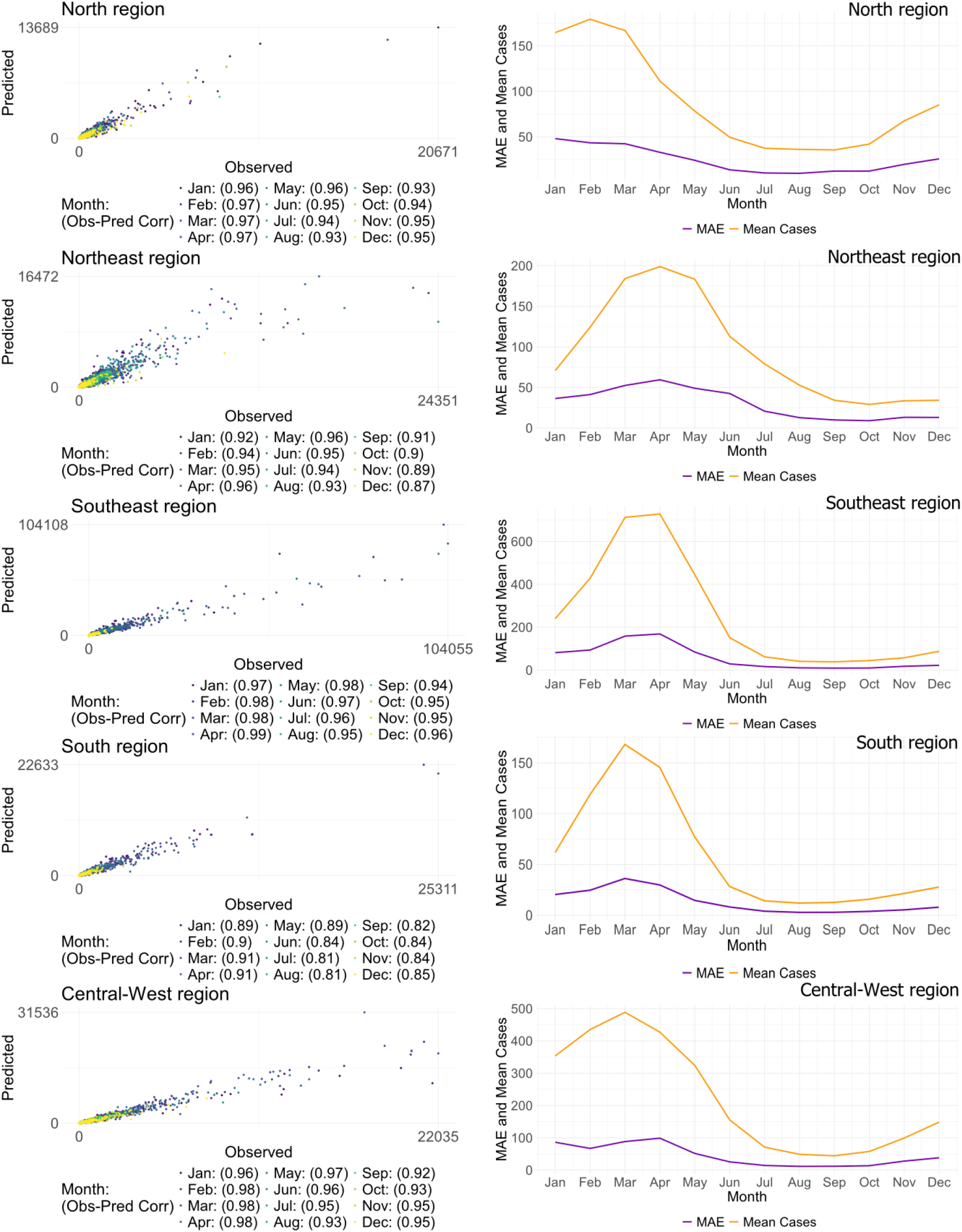
Results from the predictive model cross-validation: correlation between observed and predicted cases by region and month of the year (left panel), and mean absolute error (MAE) versus the mean cases by region and month (right panel)

For the future projections of dengue incidence rates, we used the UKESM1-0-LL model [22], a high-complexity climate model that simulates various atmospheric, oceanic, terrestrial, and biogeochemical processes. We present three different climate change scenarios described by the Shared Socioeconomic Pathways (SSP): SSP126 (temperature increase of 1.18 °C/100 yr), SSP370 (temperature increase of 3.6 °C/100 yr), and SSP585 (temperature increase of 4.4 °C/100 yr).

These scenarios represent low, high, and very high greenhouse gas emissions, respectively [22–25]. The description of the climatic variables minimum temperature (lag 1 month) and precipitation (lag 2 months) for the period 2021-2040 and 2041-2060 are presented in Figure 3. It can be observed that there is a clear increase in minimum temperatures in future scenarios compared to the historical period of 2000-2021, with the last period showing the highest increase. There is also generally little difference between scenarios, with the greatest difference seen in the last period where scenario SSP585 has the highest temperature values, followed by SSP370, and finally SSP126, across all regions. Precipitation generally remains constant, with a slight visual increase in peaks in the northeast, southeast, and central-west regions, and a slight visual decrease in other regions.

**Figure 3:**
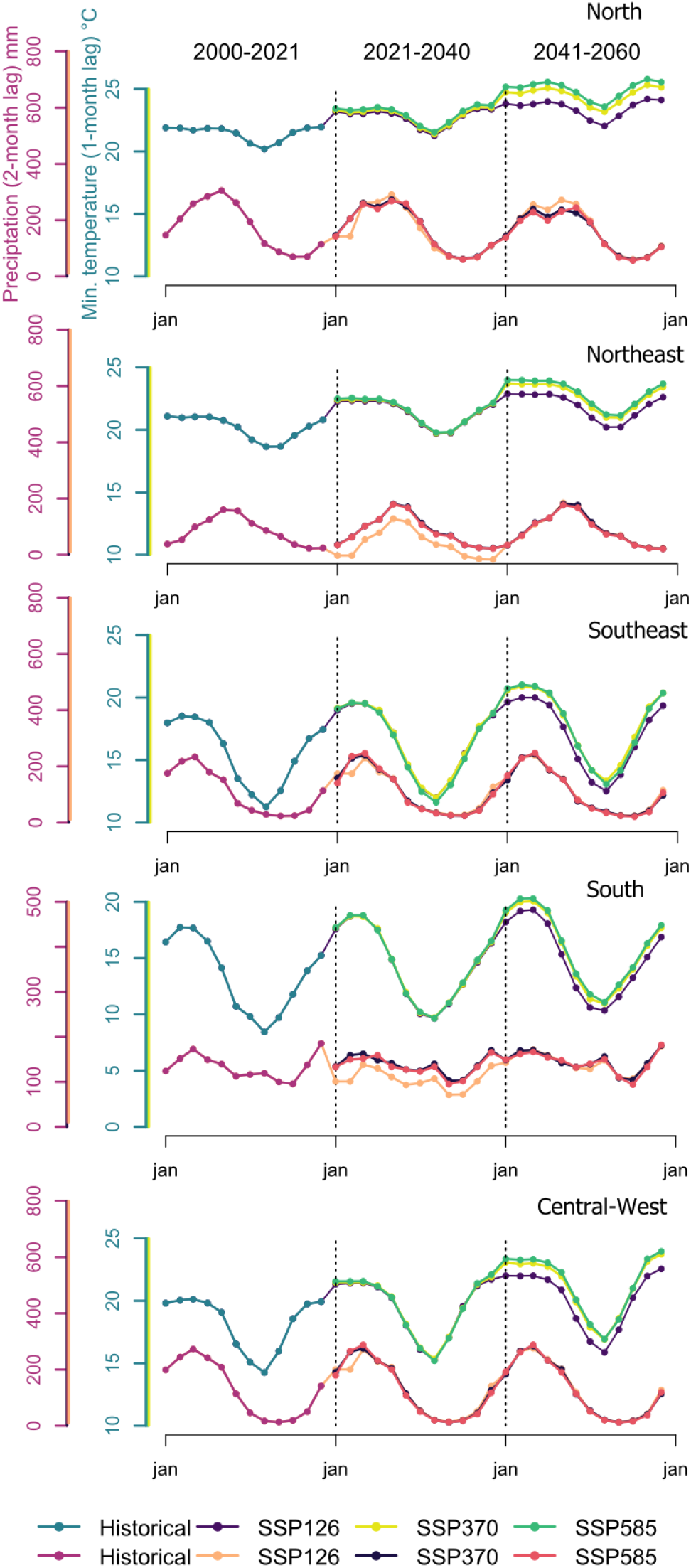
Distribution of precipitation (purple y-axis) and minimum temperature (green y-axis) variables by month: historical period (2000-2021) and future projections (2021-2040 and 2041-2060) across three18 different SSPs scenarios

The average population for the period 2000-2021 was approximately 195 million, rising to average values of 220, 235, and 219 million for scenarios SSP126, SSP370, and SSP585 respectively from 2021-2040, and reaching average values of 219, 258, and 217 million in the respective three scenarios considered for 2041-2060. Similarly at regional level, scenario SSP370 had the highest population increase, with scenarios SSP126 and SSP585 having values close to each other.

Figure 4 presents the posterior predictive mean of the projections for the month of March, chosen due to the highest dengue incidence. This figure includes the historical years 2000, 2016 (peak year), and 2021, as well as the predicted periods 2021-2040 and 2041-2060, for the three scenarios SSP126, SSP370, and SSP585.

**Figure 4:**
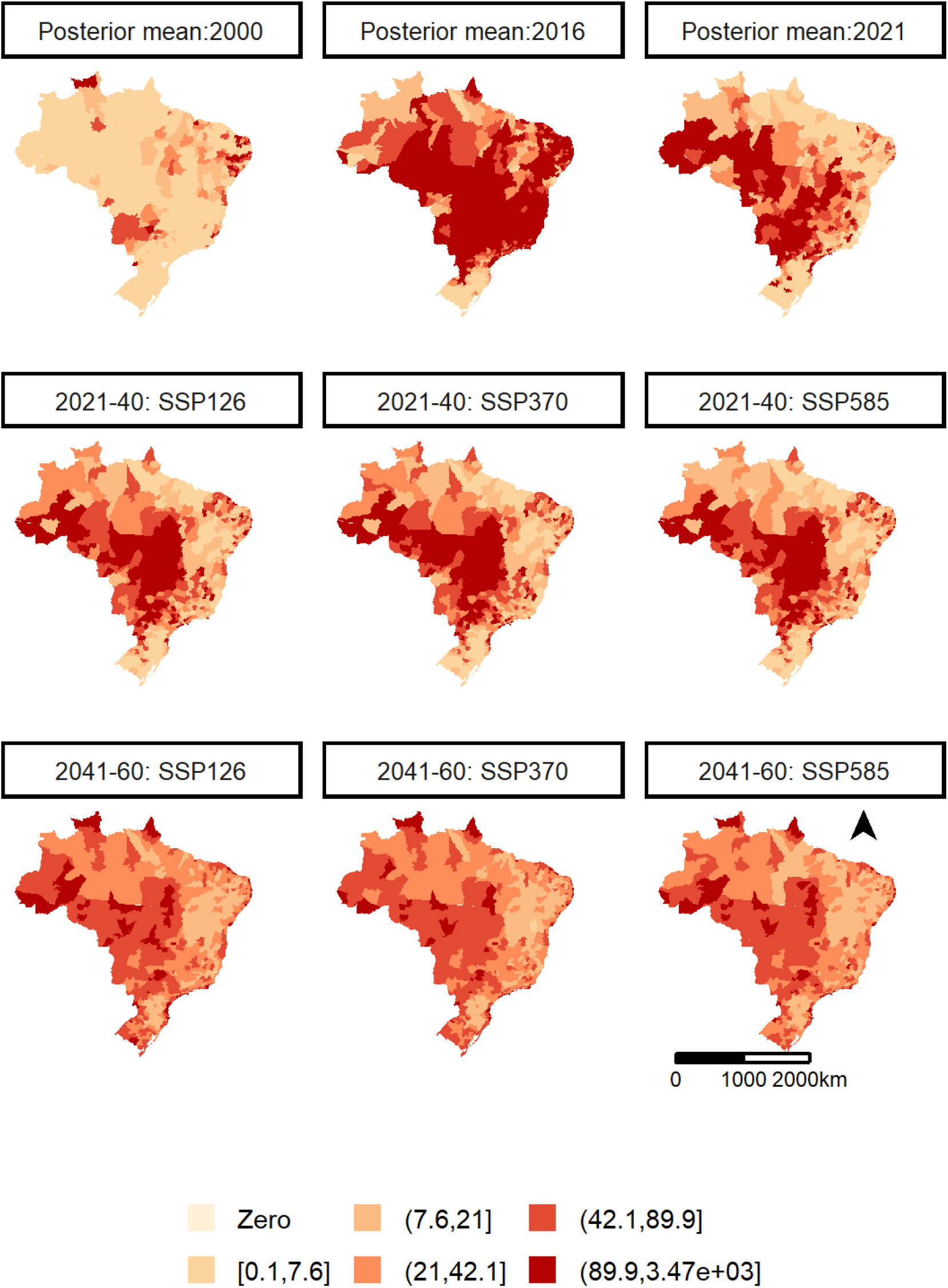
Spatial distribution of the posterior mean of dengue incidence rate for the month of March: years 2000, 2016, and 2021, and future projections 2021-2040, 2041-2060, across three different scenarios.

It is worth mentioning that, the north, central, northeast and southeast regions show a considerable geographic variability in the predictions of incidence rates, with microregions presenting increase, reduction and maintenance in their incidence patterns. Meanwhile, there is a clear increase in the posterior mean of dengue incidence rate in the south region of the country, which historically have shown low incidence scenarios. A clearer spatial visualization of the posterior means of dengue incidence for March in this region can be seen in detail in Figure 5 (panel A), for the years 2000, 2016, and 2021, and predicted values for future periods and scenarios.

**Figure 5:**
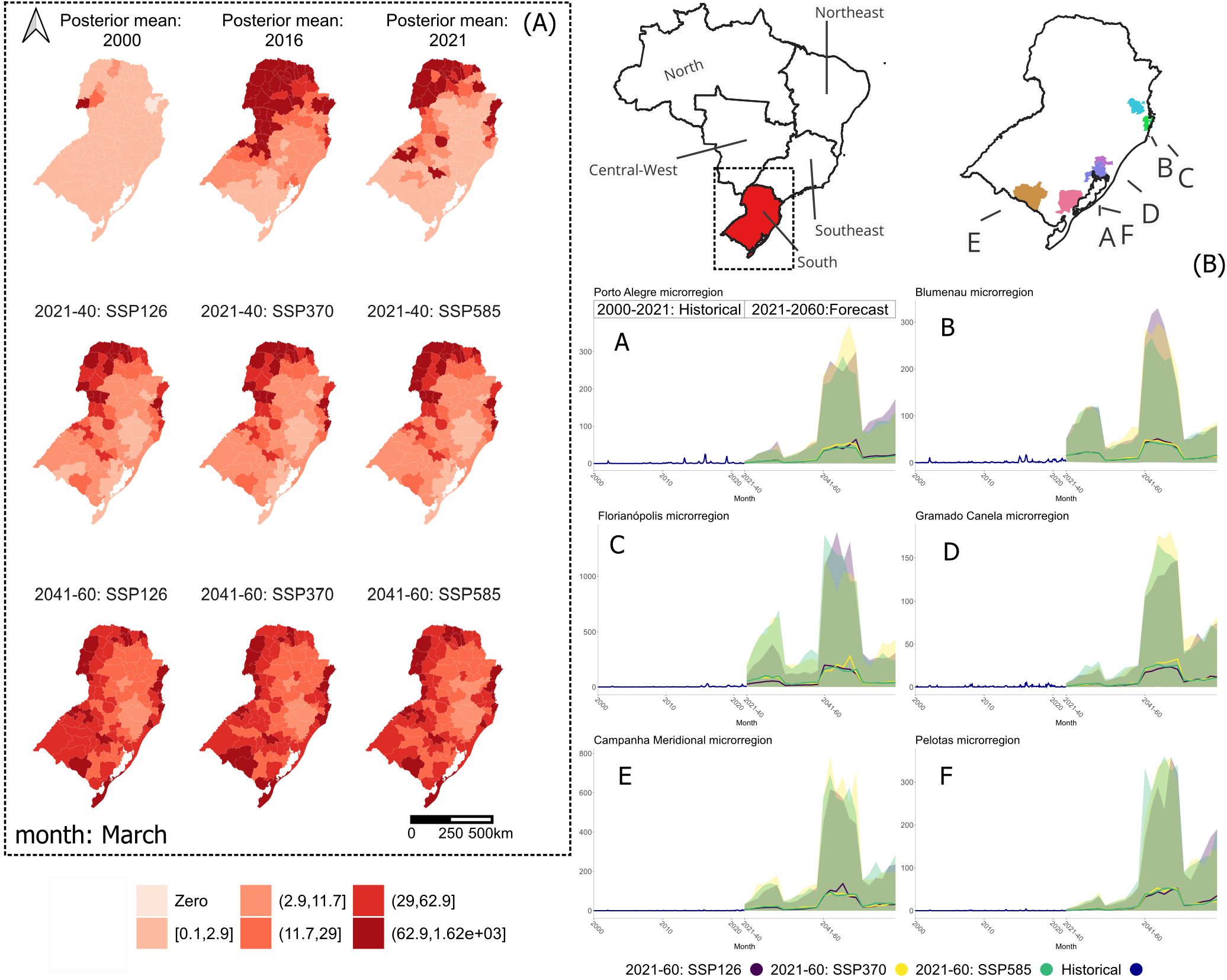
Spatial distribution of the posterior mean of dengue incidence rate for the month of March: years 2000, 2016, and 2021, and future projections 2021-2040, 2041-2060, across three different scenarios. South region of Brazil, with emphasis on temporal projections for selected microregions

Figure 5 (panel B) also indicates the temporal change of dengue incidence (posterior mean and 95% CI) for six selected microregions in the South region, chosen as those where the model predicted a considerable increase compared to the historical series.

## Discussion

This study has presented probabilistic estimates of dengue incidence in Brazil, based on climatic, demographic and socio-economic factors and, to the best of our knowledge, is the first to provide projected future scenarios for the periods 2021-2040 and 2041-2060, considering different greenhouse gas emission scenarios at microregion level.

The other only study regarding Brazil and greenhouse gas emission scenarios, by Colon-Gonzalez et al. [26], made an important contribution by estimating the increase in dengue cases under different global warming scenarios across Latin America for the years 2050 and 2100, using historical data from 1961–1990 as a baseline and considering three climatic variables. Compared to it, we believe our study brings a new perspective, as it is entirely focused on Brazil, uses more up-to-date data, incorporates additional factors beyond climatic variables in the model, and makes predictions closer to the present. By doing so, we can establish possible patterns for different Brazilian microregions, indicating not only areas with expected increases but also some with stability or even reductions in dengue cases.

As far as we know and according to the systematic review by Xu et al. [27], other studies that have aimed to predict dengue under different greenhouse gas emission scenarios commonly used the years 1961-1990 as the baseline period and 2050 as the projection period. These studies covered areas including Nepal, China, the US, Korea, 10 European cities, Australia, and globally [27]. Additionally, Colon-Gonzalez et al. [28] predicted dengue across Southeast Asia, using data from 2000–2017 and making predictions for 2050 and 2080. Recently, Wang et al. [29], used the baseline period of 2012-2020, to predict conditions for the 2030s to 2090s in South and Southeast Asia.

In our study, we identified a lagged and nonlinear behaviour of minimum temperature (modelled with a 1-month lag), which showed a negative relationship with dengue incidence until reaching 14.4°C, peaking at 21.2°C. This result indicates that very low temperatures have an inverse association with disease incidence, while at higher temperatures we found a positive association with dengue incidence [30]. This result is in line with the current evidence, showing that temperature influences the spread of dengue. Laboratory tests indicate that the temperature for mosquito population development is 13°C, with a maximum threshold of 36°C [31, 32]. Mordecai et al. [33] found that, in a more realistic scenario where daily temperatures varied by 8°C, the minimum and maximum temperatures for transmission were 13.5°C and 34.2°C, respectively. The systematic review by Abdullah et al. [34], on the other hand, found that the association between dengue cases and minimum temperature ranges from 6.5°C to 21.4°C, discussing that *Ae. aegypti* has a higher low-temperature tolerance compared to *Ae. albopictus*. Therefore, although studies report varying minimum temperature values linked to dengue cases, our analysis of monthly data from Brazilian microregions aligns with existing literature.

As anticipated, the correlation between precipitation and dengue incidence proved to be positive, with a two-month lag, which is consistent with the literature [30]. This means that as the amount of precipitation increases, the incidence of dengue also tends to rise. One of the phases of the vector’s reproduction occurs in aquatic environments, meaning that mosquitoes lay their eggs in places with standing water. Therefore, more rain results in more breeding sites and, consequently, an increase in the mosquito population. This leads to a higher probability of dengue transmission [35].

Since there is a time interval required for the eggs to hatch and for adult mosquitoes to develop and start transmitting the virus, it is expected that dengue incidence will increase about two months after a rainy period [30]. This time frame allows mosquitoes to complete their life cycle and begin infecting people, resulting in an increase in dengue cases. Additionally, rainy periods often bring higher temperatures and humidity, favoring mosquito activity [36].

However, it is important to note, that precipitation alone may not conclusively determine vector presence. Even in arid environments, transmission can persist, as communities often resort to rainwater harvesting during such periods. Furthermore, the vector’s eggs are typically found on container walls, above the waterline, and they can remain viable for over a year without water contact, underscoring that the absence of water does not necessarily eradicate the vector population [31].

As expected, we found a positive relationship with the NDWI, in line with other studies. This suggests increased water availability, potentially leading to higher mosquito larval reproduction and survival in stagnant water containers such as tires, water storage containers, plant pots, and other objects that collect water [37]. Therefore, a positive NDWI may indicate areas prone to *Ae. aegypti* mosquito breeding, thereby posing a higher risk of dengue transmission. Additionally, NDWI can aid in identifying flood-prone areas, which may further increase breeding sites for vector mosquitoes. Thus, the positive association between NDWI and dengue underscores the importance of water presence in predicting and controlling dengue incidence [38, 39].

The percentage of urban infrastructure also showed a positive association with dengue incidence. This result is expected since *Ae. aegypti*, the primary vector, is predominantly urbanized, with a preference for oviposition in artificial containers. Urbanized areas, therefore, present ample opportunities for breeding sites for the vector, ranging from trash heaps and cemeteries to residential areas, where 75% of breeding sites are located, according to the Ministry of Health [40]. Additionally, the proximity of people in urban areas provides further sustenance to the vectors [41].

Our study found an inverse relationship between dengue cases and elevation, which is expected due to the lower temperatures [42, 43].

Regarding the deprivation index, our study reveals that higher incidences of dengue are associated with microregions characterized by lower levels of deprivation. Despite these findings contrasting with some studies that observed a positive relationship, the association between dengue and socioeconomic conditions is considered controversial, and the disease affects people across a wide range of socioeconomic profiles. Mondini and Chiaravalloti Neto [44] highlighted this controversy in their evaluation of the relationship between dengue risk and socioeconomic conditions in a Brazilian area, emphasizing its potential variability across different municipalities.

Therefore, it is crucial to interpret these results cautiously due to the study’s geographical scope, encompassing microregions composed of clusters of municipalities. Additionally, the most recent available socioeconomic data stem from the 2010 census. This temporal gap underscores the need for updated and localized socioeconomic assessments to better understand the nuanced dynamics of dengue transmission in diverse socioecological contexts.

Lastly, we found a positive association with the summer and autumn seasons compared to winter, and no significant association with spring. This result is expected for Brazil, as it is well known that dengue follows a seasonal pattern, with higher prevalence in summer and lower in winter [45].

It is well-established that climatic variables significantly impact the severity and spread of the disease. Temperature plays a dual role by hastening the vector’s life cycle and enhancing its ability to transmit the virus, while also shortening the virus’s extrinsic incubation period of the vector, thus expediting transmission [46, 47].

Consequently, it is expected that climate changes can profoundly alter both the distribution and severity of dengue incidence, potentially amplifying or mitigating its impact [31, 35]. Substantiating this, our study shows a pattern of increased dengue incidence in microregions with historically low cases, while observing either maintenance or even a decrease in areas with previously high case rates.

Our findings place important emphasis on the south region of Brazil. This region is characterized by well-defined climatic seasons, featuring lower temperatures compared to other regions of the country, factors that hinder mosquito proliferation. Consequently, this region exhibits the lowest incidence rates for the historical period [48].

Although the first case of dengue in Brazil was reported as early as the 1980s, after the reintroduction of *Ae. aegypti*, the initial case in the south region resulted from the introduction of serotype 2 in 1995. Following this, serotype 1 was introduced in 1999, serotype 3 in 2000, and serotype 4 in 2011 [49].

Soek et al. [31] analyzed climatic variables based on future scenarios predicted by the IPCC, although they did not directly associate these variables with either vector presence or disease cases. The authors concluded that these projections indicate a high susceptibility to the expansion of vector coverage areas and prolongation of periods of high dengue transmission due to the extended time within optimal conditions for vector reproduction.

Our study corroborated this hypothesis, demonstrating through robust statistical modeling that there is a projected increase in dengue incidence in this region. We found that several microregions in the south region, previously considered non-endemic, can become public health emergencies.

It is worth noting that recently, a state in the South region of the country faced a calamity event attributed to climate change. Specifically, the state of Rio Grande do Sul experienced flooding due to an extreme weather event of heavy rainfalls, affecting over one million people. This flood event is just one example of the impacts of climate change, which have the potential to exacerbate future health-related issues, such as the spread of dengue.

Climate change is already considered one of the greatest threats to life, especially for vulnerable populations. Bashir [50] underlined that climate change increases the burden on the healthcare system and has economic as well as mortality consequences. This is particularly concerning as it disproportionately affects the vulnerable population and contributes to increasing inequality.

Barcellos et al. [51] stated that climate change and thermal anomalies have played a significant role in the recent progression of dengue in Brazil. Between 2000 and 2014, they observed a notable increase in the frequency of temperature anomalies across various regions of the country, which was associated with an increase in the incidence of the disease.

Another region that deserves attention is the Central-West. This region recorded the first case of serotype 1 in 1987, followed by serotype 2 in 1991, serotype 3 in 2000, and serotype 4 in 2011 [48]. Barcellos et al. [51] discussed the increase in thermal anomalies and found that the Central-West region had the highest incidence of dengue in the country. In our study, future projections indicate that this region will continue to have the highest incidence rates.

In general, we observed that North, Northeast, Southeast and Central-West regions showed a visual geographic variability in the pattern of dengue incidence, with increase, stability or even decline for the future. The decrease that we predicted for several microregions corroborates with the study developed by Cardoso-Leite et al. [52], which predicted the environmental suitability of *Ae. aegypti* under current and 2050 climatic conditions. They found a projection of decreased vector suitability in the future for the North region of Brazil, which is likely due to temperatures surpassing the maximum viable temperature threshold for the vector.

One limitation of our study is the challenge of modeling dengue incidence in an unstable climate context. To address this limitation, we utilized robust generalized linear mixed effects models with spatio-temporal random fields to capture spatial and temporal dependencies. Additionally, we considered three different scenarios of future climate projections along with projection of population growth and we performed a rigorous cross-validation to evaluate the predictive ability of the model.

One additional limitation to consider is that the socioeconomic data are from the latest census conducted in 2010. However, this is the most current data available, as the 2022 census results have not yet been released.

Future projections of minimum temperature and precipitation used in our study are from WorldClim [53, 54], which provides monthly information for each 20-year period rather than annual data. Nevertheless, despite this limitation, the 20-year periods yield more stable statistical estimates, reducing short-term fluctuations and allowing for more consistent comparisons across microregions. WorldClim data remains an invaluable source because it is derived from terrestrial weather stations, satellite data, and global climate models, and is processed and interpolated to create high-resolution maps representing climatic conditions.

While the most recent available historical data year is 2021, the time of writing this paper, and has yet to be updated with more recent data, future studies could explore the potential of evaluating these estimates using newer data as it becomes accessible through WorldClim. Finally, future studies should also emphasize the importance of assessing the impact of vaccines, especially considering that in 2024, the public distribution of vaccines began through the Unified Health System (SUS), initially targeting younger age groups [18].

To conclude, we stress that dengue is a complex challenge in public health, persisting in causing major epidemics and spreading territorially, despite decades of efforts dedicated to its surveillance and control. Our results indicate that, according to the different greenhouse gas emission scenarios considered, the threat will persist and possibly it will change its spatial pattern in some Brazilian microregions, worsening in some areas and mitigating in others.

We hope that these results highlight the importance of cross-sector investments in surveillance and control, focusing on areas of highest risk. Additionally, we hope that our findings underscore the relevance of the consequences that climate change may have for society, including the neglected disease burden and the social and economic impacts caused by dengue.

### Subsection for Method

Brazil has an estimated population of around 203 million inhabitants (IBGE 2024) and covers a land area of 8,510,820,623 km^2^. The Gross Domestic Product (GDP) per capita was RS 31,833.50 (2017), with a birth rate of 14.16 live births per thousand inhabitants (2015). The illiteracy rate among those aged 15 years and older was 6.6 % (2019), while the infant mortality rate stood at 12.35 deaths per thousand live births (2018). The unemployment rate was 11 in the fourth quarter of 2019, and the Gini index, which measures income inequality, was 0.61 in 2010. The units of analysis for this study were the 557 microrregions of Brazil.

### Historical data: 2000 to 2021

Information on dengue cases was acquired by month and year and municipality of residence, of Notifiable Diseases Information System - Sinan, provided by the Ministry of Health, through a formal request. Subsequently, the cases were aggregated by microregion to overcome the problem of data sparsity. To achieve this, we utilized a table of the Brazilian Territorial Division, which provides the correspondence between each municipality and its respective microregion, as established by the Brazilian Institute of Geography and Statistics (IBGE).

The total precipitation and minimun temperature were obtained monthly from CRU-TS 4.06 [53] downscaled with WorldClim 2.1 [54], with a spatial resolution of 2.5 minutes (∼21 km^2^ at the equator).

In the model, it was important to consider the seasons of the year because this variable captures seasonal patterns in disease transmission and vector activity. It is known that the periods of highest disease transmission are during summer and autumn.

Precipitation is also an important variable for the disease transmission pattern. During rainy periods, suitable conditions are provided for mosquito development in breeding sites, as females lay their eggs in stagnant water, which hatch when the water level rises due to rainfall. There is an expected lag between increased precipitation, increased vector abundance, and disease emergence [35].

Socioeconomic data was derived from the Brazilian Deprivation Index (BDI), a metric designed to gauge socioeconomic status by incorporating data on income, education, and household conditions gathered during the 2010 Census by IBGE. BDI is a valuable tool for monitoring and assessing socio-economic factors’ impact on public health. This index was developed by researchers from the Center for Data Integration and Knowledge for Health (Cidacs/Fiocruz Bahia) and the University of Glasgow-Scotland, as part of the Social Policy and Health Inequalities (SPHI) project, funded by the UK’s National Institute for Health Research (NIHR). In terms of interpretation, values close to zero signify moderate deprivation, positive values indicate high deprivation, and negative values represent low deprivation. The BDI was transposed from municipalities to microregions by aggregating and weighting it based on the population size.

The urban infrastructure percentage was derived from images in the MapBiomas collection 7.1, by year and months of study, a collaborative project by various Brazilian institutions aimed at mapping land use and cover [55]. The elevation data were obtained from NASA Shuttle Radar Topography Mission (SRTM) with a resolution of 1 arc-second (approximately 30 meters).

We acquired the normalized difference water index (NDWI) for each microregion at each point in time from a series of data images from the Moderate-resolution Imaging Spectroradiometer (MODIS) surface reflectance, constructed using 16-days of Terra and Aqua MODIS data at 500 meter resolution. The NDWI is an indicator for vegetation liquid water content [55,56], and allows to differentiate water from the dry land.

### Future data: 2021 to 2040 and 2041 to 2060

The future climate variable predictions were based on documents provided by the Intergovernmental Panel on Climate Change (IPCC). The global climate model (GCM), specifically the UKESM1-0-LL model, was selected for this study due to its stable representation of a realistic climate, vegetation, and anthropogenic and natural aerosol states without requiring artificial corrections [22].

Three climate scenarios, based on the Shared Socioeconomic Pathways (SSPs), were considered to describe the evolution of greenhouse gas emissions. The SSPs (including SSP1, SSP3, and SSP5) are valuable tools for studying the potential consequences of different socioeconomic and environmental trajectories on climate change [57]. These scenarios allow re-searchers to assess the implications of various policy choices and societal pathways on global temperature rise, informing decision-making and facilitating the development of strategies to mitigate climate change and achieve sustainable development goals.

Developed by the scientific community, the SSPs provide a range of plausible socioeconomic and environmental storylines that explore alternative future trajectories of human societies and their interactions with the environment, including climate change [57].

SSP1, Sustainability, depicts a world with low challenges to mitigation and adaptation, emphasizing inclusive development, environmental stewardship, and human well-being. Improved global governance and investments in education and health reduce inequality and shift consumption towards low material growth and resource use. SSP3, Regional Rivalry, involves high challenges to both mitigation and adaptation. Nationalism and regional conflicts lead to a focus on security over development, with slow economic growth and persistent material-intensive consumption. Inequalities worsen, and environmental degradation increases due to the low priority on environmental issues. SSP5, Fossil-fueled Development, presents high challenges to mitigation but low challenges to adaptation. Driven by competitive markets and technological innovation, this scenario sees rapid economic growth and human capital development, with significant fossil fuel use. Local environmental issues are managed, and there is confidence in addressing broader social and ecological challenges, including geoengineering [58].

In detail, SSP1-2.6 (SSP126) represents a mitigation pathway with a projected global temperature increase of 1.8°C (1.3 to 2.4°C). SSP3-7.0 (SSP370) represents a high-emission pathway with a projected increase of 3.6°C (2.8 to 4.6°C). SSP5-8.5 (SSP585) represents a very high-emission pathway with a projected increase of 4.4°C (3.3 to 5.7°C) [57].

The climate data (minimum temperature and precipitation) for these three scenarios were provided by WorldClim 2.1 for two time periods: 2021-2040 and 2041-2060, with 2.5 minutes of arc (∼21 km^2^) resolution [54].

Forecasts for 2021-2040 and 2041-2060 from WorldClim are based on monthly averages for this 20-year period, as there are no yearly data available. We considered to be appropriate to use 2021 as the most recent year in our historical data and to employ the 2021-2040 forecasts as a representative monthly average for the future. Additionally, predicting the 20-year average rather than specific yearly values can offer a more stable perspective and be less susceptible to annual variations, allowing for more robust planning.

In addition to climate variables, population data for the future was also used, with a resolution of 30 seconds (∼ 1km) based on the selected SSPs scenarios [59].

### Statistical model

We employed a Bayesian hierarchical model to project the future evolution of dengue incidence to 2060 for each Brazilian microregion under three climate scenarios (SSP126, SSP370, and SSP585).

Let *Y*_*it*_ and *N*_*it*_ be the number of dengue cases and total population (per 100,000 inhabitants) in microre-gion *i* ∈ (1, …, *n*_*t*_) during month *t* ∈ (1, …, *T*), respectively. Being the outcome variable, *Y*_*it*_, a count, we specified a Poisson distribution. Thus, the first level of the Bayesian hierarchical model has a Poisson log-linear specification given by:

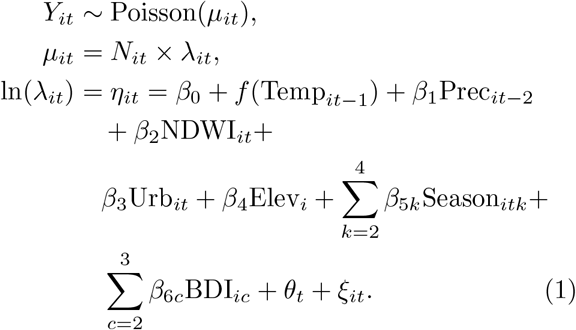

Here, *µ*_*it*_ represents the Poisson mean, which is a product of *N*_*it*_, the total number of populations in each microregion *i* at time *t*, and *λ*_*it*_, the underlying unknown dengue incidence rate in each microregion *i* at time *t*. The log-dengue incidence rate is modelled in function of *p* known climatic, environmental and socioeconomic covariates, related to microregion *i* and month *t*, including the intercept term, *β*_0_, and two random effects capturing a residual unstructured temporal component, *θ*_*t*_, and spatiotemporal correlation, *ξ*_*it*_. In detail, *f* (Temp_*it*−1_) capture nonlinearity in the effect of temperature [Temp] lagged of one month, *β*_1_ is the coefficient associated with precipitation [Prec] lagged of two months, *β*_2_, *β*_3_, *β*_4_ are the coefficient associated to the linear covariates NDWI, urbanization [Urb] and elevation [Elev] respectively, *β*_5*k*_ is the regression coefficient associated with calendar season [*Season*] (reference category: winter), and *β*_6*c*_ is the regression coefficient associated with BDI (reference: low BDI level, i.e., most deprived).

Additionally, to capture the unstructured temporal variation over months and years, *θ*_*t*_ was modeled using a Gaussian white noise process. This allowed capturing the temporal variation unexplained by the model and structured effects. The space-time latent effect *ξ*_*it*_ is modelled by a zero-mean Gaussian Markov random field with covariance matrix that implies that within each time point the microregion are linked through an intrinsic conditional autoregressive prior [60], which incorporates a spatial dependency structure among the microregions represented by the graph of the neighborhood matrix [61], while over time (i.e., between time points) the process evolves dynamically according to a first order autoregressive process with coefficient *ρ <* 1 [62, 63].

The regression parameters were modelled assuming independent weakly informative zero-mean Gaussian prior distributions with variance of 1000, while penalising complexity (PC) priors [64] were used for the hyperparameters of the random walk structures and the autoregressive process coefficient, which are expressed through probability statements. In particular, we specified that the probability for the standard deviations of the first order random walk models of being greater than 1 is small, and is equal to 0.01, and that the probability for the *ρ* parameter to be larger than 0 is large being 0.9. To model the nonlinear relationship between minimum temperature, lagged by one month, and the response, we binned the temperature values into *m* groups, using equidistant quantiles in the probability space and we specified a first order random walk prior over the *m* groups. This parameterization posses the necessary flexibility to accommodate complex patterns of variation. Specifically, we assumed:

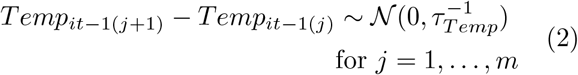

where *τ*_*Temp*_ is the precision (i.e., the inverse of the variance). We imposed a sum to zero constraints to make it identifiable, and for easier interpretation we scaled the model to have an average variance equal to 1.

### Future projections

Spatio-temporal projections for the period 2021-2040 and 2041-2060, under climate scenarios SSP126, SSP370, and SSP585, were obtained by sampling from the posterior predictive distribution (PPD) gomez2020bayesian, orozco2023scalable. Broadly speaking, if 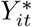 represents the new observation *I* at a future time point *t*^*^, the PPD is defined as: 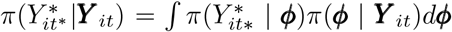 where ***Y*** _*it*_ ={*Y*_*i*1_, *Y*_*i*2_, …, *Y*_*iT*_} and ***ϕ*** indicate all the model parameters. As described in the previous section, in our study we built a multilevel statistical model including a number of predictors. Thus, by letting **X**_*it*_ to represent collectively the model predictors described in eq. [1] for the years 2000-2021 and letting 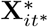 to be the future predictors for the period 2021-2040 and 2041-2060 under the different climate change scenarios, the PPD was defined as:

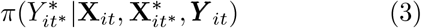

Specifically, by fitting the model in eq. [1], we obtained the approximated joint posterior marginal of ***η*** and ***ϕ***, then we sampled from it by using 1000 draws, then we generated the future projected dengue disease cases from a Poisson distribution with mean 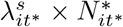, for *s* = 1, …, *S* = 1000, where 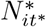 are the new future population estimates for the period 2021-2040 and 2041-2060.

### Model assessment and cross-validation

Prior to develop the statistical model, we performed a number of descriptive analyses in order to learn about relationship between the data. This included using the VIF (Variance Inflation Factor) to check for collinearity among the variables, retaining only those with a VIF value below 3. Then, the models were built from the simplest to the most complex following a forward selection procedure, testing also for nonlinearity and lagged effects of the climate variables [65].

Since the prediction objective was to use the historical series to forecast subsequent periods, in the *Time-wise Holdout* procedure, models were run excluding last year of the series (2021) as validation data and using the historical series to train the model, similarly to Oliveira and Colleagues [66].

Moreover, an additional validation performed on the best model to verify the predictive capacity of the model was thoroughly assessed through a robust Spatio-temporal Contiguous Block Cross-Validation procedure [66]. This process involved systematically removing data from the microregions of a specific geographic area and a specific month from the model. The evaluation metrics used to determine the model’s performance included the coverage of the 95% prediction intervals (obtained by counting how many of the values in the validation set fall in their corresponding prediction interval), the mean absolute error (MAE), and the Pearson’s correlation between observed and predicted count of dengue cases.

To qualify as a good predictive model, coverage equal or close to the nominal level (that is, 95% for a 95% prediction interval) is expected, indicating that the model reliably captures the variability in the data, as the model is able to predict new data with a precision consistent with what was observed in the training data. Additionally, there should be a high correlation between the predicted and observed values, suggesting that the model accurately captures the underlying patterns and trends present in the observed data, reflecting its predictive strength. Furthermore, MAE should be lower than the mean value of the observed cases, demonstrating that the model performs better than a basic model that might sim-ply predict the average value for all instances [28]. A lower MAE signifies that the model’s predictions are closer to the actual values, underscoring its accuracy and effectiveness.

The cross-validation in space and time adds a significant layer of rigor to the evaluation process. It ensures that the model is not only capable of making accurate predictions based on the existing data but also robust enough to generalize well across different spatial and temporal contexts. This comprehensive validation approach is crucial for confirming the reliability and generalizability of the model in various real-world scenarios [66].

The analyses were conducted using the statistical software R version 4.2.1 (2022-06-23), while computation was performed using the Integrated Nested Laplace Approximations (INLA) algorithm [67, 68].

## Data Availability

All data produced in the present study are available upon reasonable request to the authors

## Ethics Committee approval

This study received approval from the Ethics Committee of the School of Public Health, University of São Paulo (CAAE: 51292221.6.0000.542)

## Acknowledgment

PMMB, RGSP and FCN were supported by FAPESP, process numbers: 2020/12371-7 and 2021/11721-7 (PMMB); 2021/10212-1 and 2024/01315-0 (RGSP); and 2020/01596-8 (FCN). FCN was also supported by CNPq (PQ-1C, 304391/2022-0). MP was supported by the Wellcome Trust Seed Award in Science 217362 Z 19 Z. MP and MB were also partly supported by the MRC Centre for Environment and Health, which is funded by the Medical Research Council (MR/S019669/1, 2019–2024)

## Author Contributions

Conceptualization: PMMB, MP, FCN; Data curation: PMMB, RGSP, AO; Data analysis: PMMB, MP, MB, FCN; Investigation and methodology: PMMB, MP, MB, FCN; Writing: All authors; Review: All authors; Supervision: FCN, MP.

## Conflicts of Interest

There are no conflicts of interest to declare.

## Notes

### Competing Interest Statement

The authors have declared no competing interest.

### Author Declarations

This study received approval from the Ethics Committee of the School of Public Health, University of Sao Paulo (CAAE: 51292221.6.0000.542). All data used are secondary and have been processed and presented only in an aggregated form by Brazilian microregions.

